# Short-term variability of chronic musculoskeletal pain

**DOI:** 10.1101/2025.01.12.25320413

**Authors:** Xuanci Zheng, Swati Rajwal, Carl Ashworth, Sharon Yuen Shan Ho, Ben Seymour, Nicholas Shenker, Flavia Mancini

## Abstract

**BACKGROUND:** Chronic musculoskeletal (MSK) pain can be characterized by its temporal variability and evolution, affecting both pain management and treatment outcomes. While pain variability is traditionally studied over long timescales (e.g. days or weeks), few studies have explored short-term fluctuations (e.g. minutes to seconds) and their clinical relevance. This study investigated the short-term variability of chronic musculoskeletal pain across consecutive days, examining whether these fluctuations are stable, exhibit consistent temporal patterns, and relate to clinical severity. We also explored whether individuals with chronic MSK pain could predict their pain intensity on the following day, suggesting an ability to learn about their pain’s levels.

**METHODS:** Eighty-one participants with chronic MSK pain to the back, neck, leg or arm rated their pain continuously over two days, using a smartphone-based app.

**FINDINGS:** Results indicated that pain ratings were stable and exhibited consistent temporal patterns across days, with a temporally correlated structure. High mean pain levels were associated with lower variability, possibly reflecting a stabilized pain state. Short-term pain variability negatively correlated with clinical severity, indicating that greater variability is linked to milder pain.

**IMPLICATIONS:** These findings highlight the importance of short-term variability as a distinct and clinically relevant feature of chronic MSK pain, with implications for personalized pain management strategies.

## 1 INTRODUCTION

One of the most important clinical features of chronic ongoing pain is its temporal variability and evolution, as it affects pain management and treatment. Typically, the temporal evolution of pain is measured on fairly long time scales, over multiple days or weeks [1–3]. However, very few studies considered how pain varies over short timescales, such as minutes or seconds.

The transmission of endogenous noxious signals to the human brain is characterized by a volatile and autocorrelated pattern [4, 5]. Continuous ratings of spontaneous pain have been shown to exhibit fractal properties, i.e. a power-law relationship between variability and time-scale length. Such properties differed between types of chronic pain (for example, back pain and postherpetic neuropathy), and between thermal stimulation scores or imagined pain [5]. Importantly, short-range temporal variability may reflect how well pain is endogenously regulated. For example, pain fluctuations are associated with changes in brainstem activity, in individuals with chronic neuropathic pain [6]. To complicate matters, endogenous regulation of pain may be dysfunctional in chronic pain [7]. In support of this view, medium-term pain fluctuations (over 2-4 days) have been found to be more severe and frequent in people with high-impact vs. low-impact temporomandibular pain [8].

Here, we investigate whether the short-term variability of ongoing chronic musculoskeletal pain is stable over consecutive days, exhibits consistent temporal patterns, and is associated with poor clinical outcomes. We also explored whether people with chronic pain are capable of predicting how much pain they expect to perceive the next day, as this would suggest that they have learned some information about the temporal evolution of their pain.

To these purposes, we conducted an online, smartphone-based study where participants with chronic musculoskeletal pain to the back, leg, neck, or arm continuously rated their pain, over two consecutive days. After rating pain on the first day, they also predicted its intensity for the next day and rated their confidence in this prediction. Through this approach, our objective was to discover whether short-term pain variability is not only a distinctive characteristic of chronic musculoskeletal pain but also potentially an indicator of the stability and predictability of pain experiences, shedding light on mechanisms that could improve individualized pain management and intervention strategies.

## 2 METHODS

### 2.1 Participants and screening procedures

We recruited 200 English-speaking participants online through the Prolific platform [9] to complete a health screening questionnaire for the evaluation of eligibility. Before the screening, all participants provided their digital informed consent, adhering to procedures approved by the Department of Engineering, Ethics Committee of the University of Cambridge. In order to ensure complete anonymity, the online questionnaires and tasks did not require participants to provide their names or contact details. The initial screening survey included a brief health questionnaire (Appendix A.1), focusing on medical history, along with the standard Musculoskeletal Health Questionnaire (MSK-HQ) [10]. MSK-HQ (scores between 0 and 56) enables individuals with musculoskeletal conditions to report their symptoms and quality of life in a standardized way.

To be included in the experiment, participants had to meet the following criteria: (a) having provided digital consent; (b) experiencing pain in the back, leg, neck, or arm for more than 6 months, (c) having passed an attention check question, and (d) not having any neurological, psychiatric, or developmental disorders. Based on these criteria, 123 participants were invited to take part in the two-day online experiment detailed in Section 2.2. Of these, 81 participants successfully completed the experiment on the first day and 41 participants completed both days. Figure S1 presents a flow chart illustrating the inclusion and exclusion of study participants.

Among the 81 participants who completed Day 1, there were 58 females and 23 males, with an average age of 45.6 years (± 11.7), ranging from 22 to 65 years. These participants had experienced chronic pain for an average of 9.7 years (± 6.7) and scored 36.6 (± 9.0) on the MSK-HQ scale. Among the 41 participants who completed both days of the experiment, 32 were females and 9 were males, with an average age of 46.4 years (± 11.0). These participants had been experiencing chronic pain for an average of 9.8 years (± 6.6) and scored 36.2 (± 10.9) on the MSK-HQ scale.

### 2.2 Experiment Protocol

The entire experiment was conducted online, on smartphones, over the course of two days. On each day, participants were invited to engage in a continuous rating task to assess their pain intensity using an online application developed with the open-source software package PsychoPy [11]. The application was hosted on Pavlovia [12]. At the start of the study, participants were provided with detailed instructions and required to complete a short practice run to ensure they understood how the application worked. Once this was completed successfully, the participants continuously rated their pain for approximately five minutes on a vertical scale ranging from “Least Pain” to “Most Pain”.

The interface of the application and the timeline of the experiment are depicted in Figure 1. During the experiment, two attention tests were included to assess the participants’ level of engagement with the task (tapping on a star briefly appearing on the screen), at a random time within the 90-120 seconds range. The attention checks interrupted the continuous rating process, which had an average sampling rate of 55 Hz, giving rise to 3 trials of variable length. After completing the continuous pain rating, participants were asked to estimate how much they expected their pain level to be the next day, at a similar time of day, using a visual analog scale from “Least Pain” to “Most Pain”. They also indicated their level of confidence in this prediction on a scale from “Unsure” (Not confident) to “Sure” (Very Confident). Although the VAS scale’s may have non-linear properties [13, 14], minor differences between paper and mobile assessments are not clinically significant [15].

**Fig. 1.**
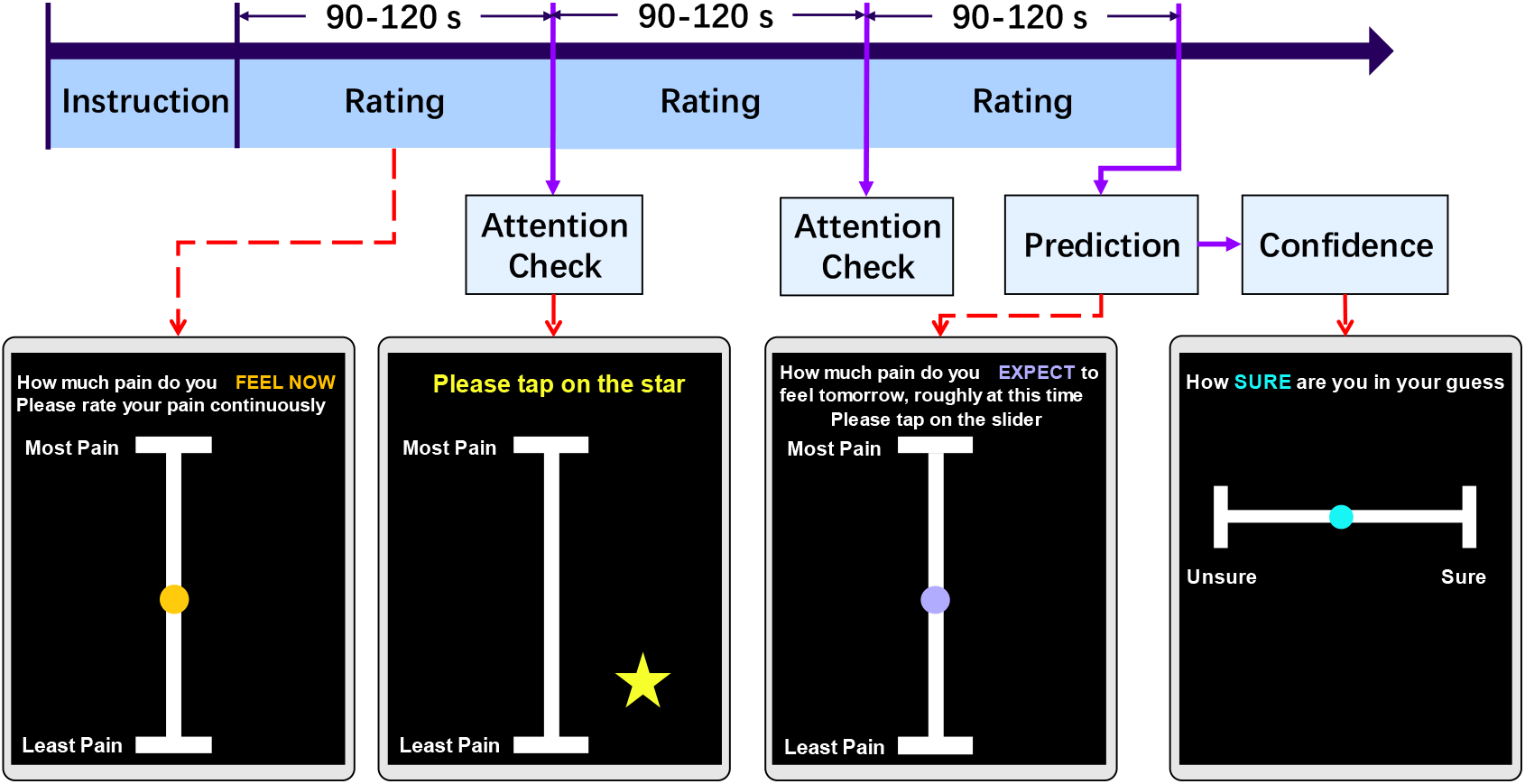
Experiment task design. Each day, participants engaged in an online pain rating task. Following instructions, volunteers were guided to continuously maintain their finger on a slider and rating their pain intensity for a randomized duration of 90 to 120 seconds. During the task, a randomly appearing star on the screen served as an attention check, which they tapped. This cycle repeated three times. Before finishing the task, participants were prompted to predict their pain levels at the same time the following day and express their confidence in this prediction by tapping the slider.

Finally, participants were asked to complete clinical questionnaires to provide a more complete description of their symptoms. Specifically, on day 1 of the experiment, participants completed the Brief Pain Inventory (BPI) [16], while on Day 2 of the experiment, they completed the Pain Catastrophizing Scale (PCS) [17]. Answering these questions was optional. The questionnaires were spread over two days to avoid participants from feeling overwhelmed by the volume of questions if they were presented all at once.

### 2.3 Data pre-processing

Firstly, we removed trials which were interrupted by the participants. Given that some participants did not keep their fingers on the screen for the entire duration of each rating, causing significant data ‘gaps’ (lack of rating data) within a trial, we segmented the rating data into epochs based on the presence of large gaps (more than 10 seconds). If any epoch had gaps for less than 10 seconds, the missing data were linearly interpolated. Epochs with more than 3 gaps that exceeded 10 seconds were deemed invalid and excluded. The total exclusion rate of trials was 23.8%. Following data segmentation, all ratings were resampled at a 40Hz sampling rate for consistency.

### 2.4 Analysis of variability and reliability of continuous pain rating

To describe the variability in the continuous ratings of the participants, we calculated for each day the mean, coefficient of variation (CV) (Eq. 1), and interquartile range (IQR) (Eq. 2), respectively, which indicate the intensity, variability relative to the mean intensity, and dispersion of the rating of each participant. For a full day’s rating by a participant, *SD* represents the standard deviation of the rating; *Mean* is the mean value of the rating; *Q*1 corresponds to the first quartile (25th percentile) of the data; *Q*3 corresponds to the third quartile (75th percentile) of the data:

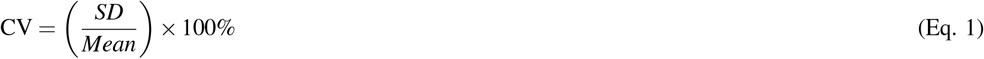

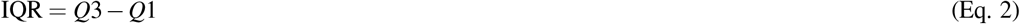

To assess the distribution of variability parameters, we generated plots for each parameter and observed that they displayed a skewed distribution. To reduce the influence of extreme values and prevent the generation of invalid results in standard statistical analyses, we applied a log transformation to reduce the skewness. Figure S2 shows the distribution of both the raw factors and the log-transformed data. The resulting distribution showed a clearer central tendency with a reduction in the spread of values.

We evaluated the test-retest reliability of the ratings on Day 1 and Day 2, by calculating the intra-class correlation (ICC) and Pearson’s correlation, corrected for multiple comparisons using a Benjamini & Hochberg correction on the False Discovery Rate (FDR). This analysis was performed to evaluate the consistency of the above variability factors obtained from the ratings of the same participant on both Day 1 and Day 2. Furthermore, we estimated the lagged autocorrelation among the pain ratings, to determine whether pain ratings at each time point were based on recent pain levels.

To investigate the clinical significance of our variability measures (CV, IQR), we correlated them with the clinical questionnaire scores (MSK-HQ, BPI severity, and PCS). The BPI severity score (out of 10) is calculated by the scores for Questions 2, 3, 4 and 5 and then dividing by 4. For consistency, we also correlated the mean pain level with MSK-HQ, BPI severity, and PCS scores. Again, the FDR was corrected for multiple comparisons. Correlations for day 1 were calculated based on participants who attended day 1, while analyses for day 2 were performed using participants who attended both days.

### 2.5 Pain prediction

After assessing the variability of the ratings, we focused on evaluating the accuracy of each participant’s pain prediction. At the end of their participation in their task on Day 1, participants were asked to predict their intensity of pain at a similar time on the following day, along with their confidence in that prediction. To gauge the precision of their predictions, we calculated the Root Mean Square Error (RMSE) between their actual pain rating values on day 2 and the predictions they made the previous day. A higher RMSE indicates a lower prediction accuracy. To quantify their prediction accuracy, we subtracted the RMSE from the highest value on the prediction slider (10). In the following equation (Eq. 3), *R*_*i*_ represents the rating data on Day 2. *Pred* refers to the pain prediction for Day 2 made on Day 1.

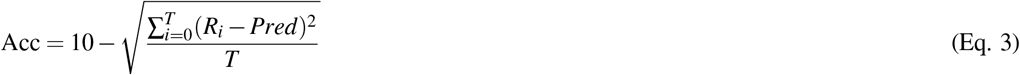

We correlated the prediction performance of participants (prediction and accuracy) with the mean and variability measures (CV and IQR).

## 3 RESULTS

### 3.1 Variability and reliability of pain ratings

The mean level of rated pain, its variability (CV) and spread (IQR) were fairly stable across the two days of testing (see Table 1).

**Table 1.** Test-retest reliability of two-day participation. Mean, CV and IQR have been calculated from ratings in Day 1 and 2. Intraclass Correlation Coefficient (ICC) and Pearson’s R were calculated between factors on Day 1 in participants who attended both days and corresponding to the same participant on Day 2.

| Factor | ICC | p-value (ICC) | Pearson’s R | p-value (Pearson’s R) |
| --- | --- | --- | --- | --- |
| Mean | 0.863 | < 0.001 | 0.823 | < 0.001 |
| CV | 0.805 | < 0.001 | 0.754 | < 0.001 |
| IQR | 0.788 | < 0.001 | 0.733 | < 0.001 |

To investigate the temporal dependencies in pain rating, we conducted autocorrelation analyses. First, we computed the autocorrelation across all lags for each participant’s individual ratings. These values were then aggregated across participants to calculate the mean and variance at each lag, providing a group-level summary of the autocorrelation structure on both days. Second, we calculated the lag-1 second autocorrelation, which measures the correlation between rating points separated by a 1-second interval, capturing short-term dependencies over a behaviorally meaningful timescale.

The visualization of the mean autocorrelation function across participants revealed a gradual decay across both days. The lag-1s autocorrelation indicated a high level of short-term dependency in the ratings (mean: 0.736 for Day 1, 0.709 for Day 2), with no significant difference between the two days (Mann-Whitney U test: U = 10855.0, p = 0.535). The group-level autocorrelation across all lags is presented in Figure 2(A), and the distribution of lag-1 second autocorrelation is shown in Figure 2(B).

**Fig. 2.**
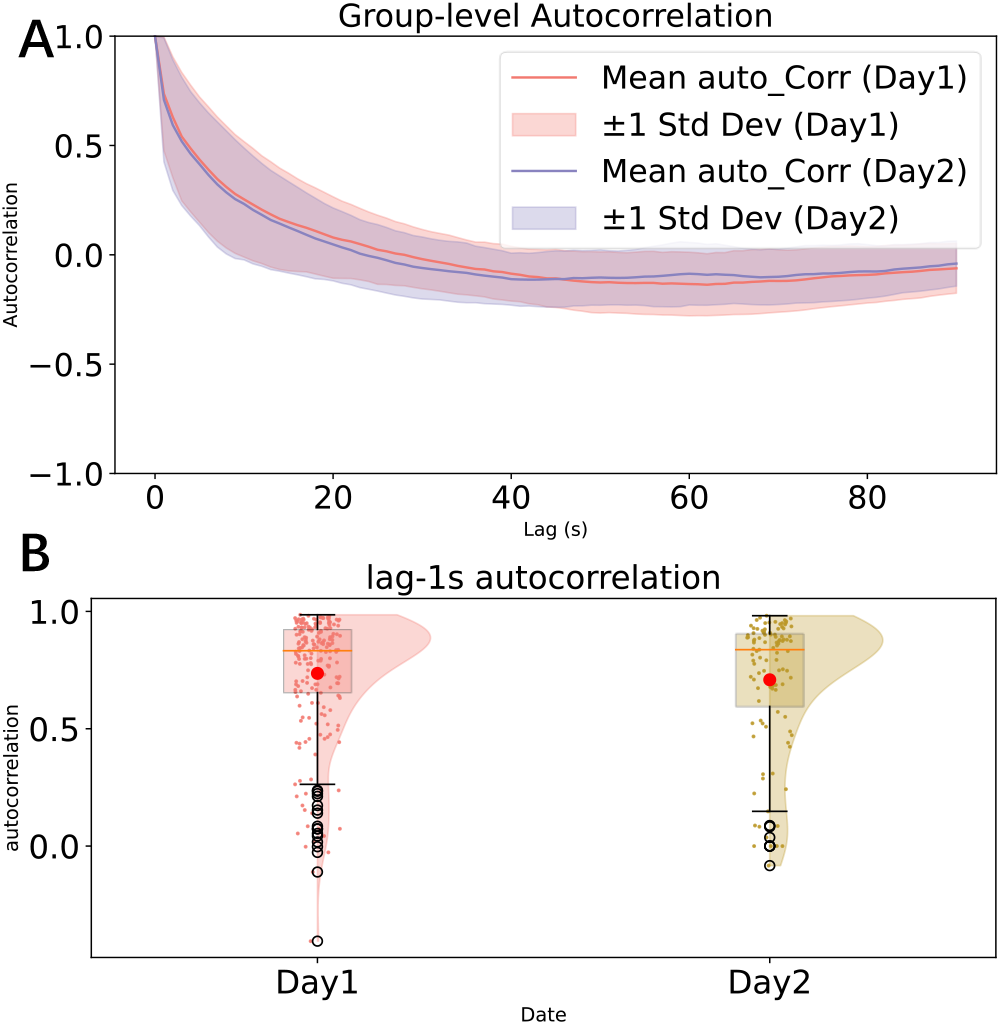
Autocorrelation analysis (A-B). **A**. autocorrelation across all lags; **B**. The distribution of lag-1s autocorrelation.

Mean pain levels were negatively correlated with CV (day 1: Pearson’s R = -0.608, corrected p *<* 0.001; day 2: R = -0.741, p *<* 0.001) and positively correlated with IQR (day 1: R = 0.382, p *<* 0.001, Figure 3; day 2: R = 0.398, p = 0.013). Hence, the more severe the intensity of the rated pain, the less variable it was, the values regressing toward their central ranges. Furthermore, as the relative variability increased, so did the data spread, but only on day 1 (CV and IQR correlation in day 1: R = 0.379, p *<* 0.001; day 2: R = 0.103, p = 0.435).

**Fig. 3.**
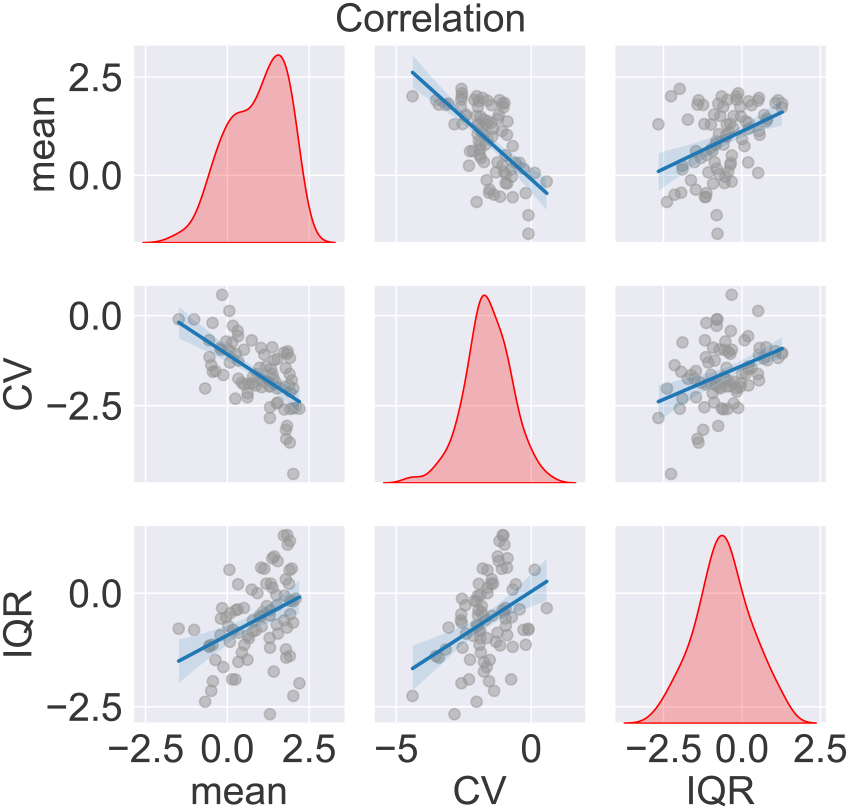
Correlation among Variability parameters (mean, CV, and IQR) on Day 1. Kernel density estimation of each factor and regression of each correlation.

### 3.2 Clinical significance

Mean pain levels and variability measures (CV, IQR) on day 1 were correlated with clinical scores to explore their clinical significance (Figure 4 and Table S1). Because the BPI was assessed on Day 1 and the PCS on Day 2, we used Day 1 data only from participants who attended both days. We found a strong negative correlation between pain variability (CV) and BPI severity and a strong positive correlation with the MSK-HQ score (n.b., the better the MSK-HQ score, the better the health status). This again indicates that the milder the pain condition, the higher its variability. The mean pain value also had a strong negative correlation with MSK-HQ and positive correlations with BPI severity and PCS.

**Fig. 4.**
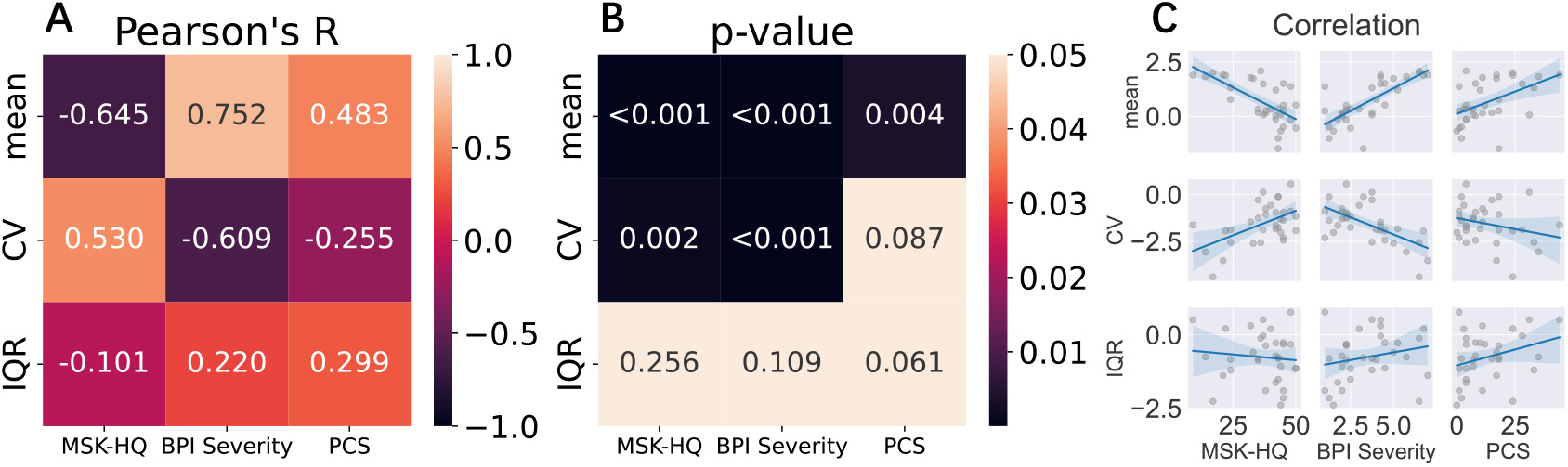
Correlation between variability measures (Day 1) and clinical outcomes (A-C). Log transformed Mean, CV, and IQR were correlated with MSK-HQ, BPI Severity, and PCS questionnaire scores. **A**: Pearson’s correlation between variability factors and clinical outcome; **B**: P-value of each correlation after multiple comparison; **C**: Scatterplots and regression lines for each pairwise relationship.

To validate these findings, the same correlation analysis was performed on day 2 data, which showed similar results (S1): CV was strongly correlated with BPI Severity and moderately correlated with MSK-HQ, while mean pain was strongly correlated with MSK-HQ, BPI Severity, and PCS. Furthermore, the spread of pain ratings around mean values (IQR) showed a moderate positive correlation with BPI severity and PCS on day 2.

### 3.3 Pain prediction

Immediately after completing the pain rating task on day 1, participants predicted how much pain they would experience the following day. The predicted pain intensity was strongly correlated with the mean pain intensity rated on day 2 and was negatively related to its variability (CV, day 2) (Figure 5 and Table S2). Furthermore, the precision of this prediction, as quantified by RMSE scores, was negatively, although weakly, correlated with the variability (CV) of pain rated on day 2, suggesting that the participants who gave the most accurate predictions may also have experienced less variable pain the following day.

**Fig. 5.**
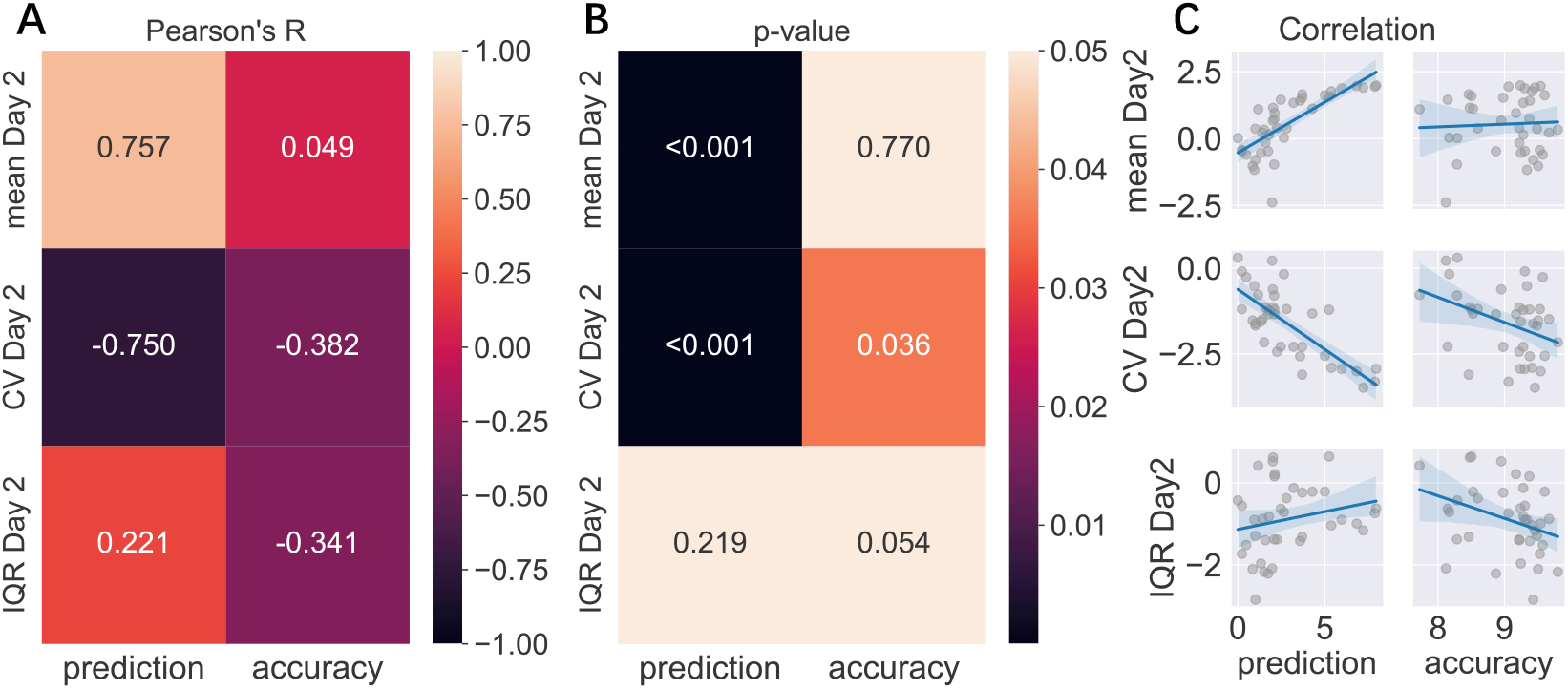
Correlation between predicted and actual pain levels and variability (A-C). Variability factors include log transformed Mean, CV, and IQR. **A**: Pearson’s correlation among predicted pain on day 1, prediction accuracy (RMSE), mean pain, and variability rated on day 2; **B**: P-value of each correlation after multiple comparisons; **C**: Regression of each correlation.

## 4 DISCUSSION

This study provides information on the short-term variability of chronic musculoskeletal pain and its implications for clinical outcomes. Our results demonstrate high test-retest reliability for mean pain levels, coefficient of variation (CV) and interquartile range (IQR). The stability of these metrics suggests that short-term fluctuations in chronic pain are not random but follow a predictable pattern over periods of several minutes, consistent with earlier work showing fractal properties in pain variability [5]. Pain variability (CV) was negatively associated with mean pain ratings, indicating that higher average pain levels correspond to less short-term variability. This may reflect more stable symptoms. In contrast, the moderate positive correlation between mean and IQR implies that higher pain levels are associated with a wider range of pain experiences within individuals.

The high lag-1s autocorrelation values observed in pain ratings suggest strong short-term dependencies, indicating that participants’ pain experience persists over brief intervals with relatively low-frequency variability. The gradual decay of the mean autocorrelation reflects a steady reduction in temporal influence over time. The consistency in autocorrelation patterns across days again highlights the stability of the temporal dynamics in pain perception.

The variability of pain ratings was also related to clinical measures. The CV of the pain ratings was negatively correlated with clinical severity measures (e.g. BPI severity) and positively correlated with health status (MSK-HQ). This suggests that people with milder pain conditions exhibit more variability in their pain experience, while those with more severe conditions have more consistent, but higher, pain levels.

There may be many different reasons for this behavior that could be explored in future studies. We do not believe that this is a simple ceiling effect because pain ratings were not so high to be considered at the ceiling level. The negative correlation between pain variability and severity might reflect stabilization of chronic pain, which in turn may depend on the sensitization of pain pathways and diminished endogenous pain inhibition [18]. This may lead to a state where pain remains too elevated and stable for too long.

Regardless of the reasons, the relationship between variability and clinical outcomes highlights the potential to use short-term pain fluctuations as a marker of pain severity. This rating assessment shows potential as a diagnostic or prognostic Patient-Reported Outcome Measure (PROM) to be incorporated into an assessment of an individual’s pain experience that could be meaningful to both clinician and patient. Future studies could assess their stability over longer time windows (weeks, months) and whether/how they are altered by different interventions. All the code required to run the online rating task is available openly on Zenodo [19].

Furthermore, our findings also shed light on the ability of individuals with chronic pain to predict their future pain levels. The strong correlation between predicted pain on day 1 and actual pain on day 2 suggests that participants could anticipate their pain levels to some extent. This aligns with the concept of temporal statistical learning, where the brain extracts patterns from autocorrelated pain signals to anticipate future pain [20, 21].

Future research could explore how interventions targeting the endogenous regulation of pain might influence short-term variability and its relationship with clinical outcomes. Additionally, investigating the neural correlates of these temporal patterns could further elucidate the mechanisms underlying pain variability and its clinical significance.

In conclusion, this study highlights the importance of short-term variability in chronic musculoskeletal pain and its potential as a predictor of clinical outcomes. Understanding these dynamics could inform more personalized and effective strategies for managing chronic pain and enhancing the quality of life of people with chronic pain. Finally, the simple continuous rating measures used here show promise in translating them into a clinically useful digital assessment that could be provided remotely.

## Data Availability

A GitHub code repository associated with the analysis in this project will be released upon the completion of peer review and acceptance of the paper. We have open-sourced the Psychopy-based pain monitoring task for use by the academic community.

https://zenodo.org/records/13754802

## 5 DATA AND CODE AVAILABILITY

A GitHub code repository associated with the analysis in this project will be released upon the completion of peer review and acceptance of the paper. We have open-sourced the Psychopy-based pain monitoring task for use by the academic community [19].

## 6 ACKNOWLEDGMENTS

The study was funded by an MRC Career Development Award (MR/T010614/1), a UKRI Advanced Pain Discovery Platform grant (MR/W027593/1), and an EPSRC research infrastructure grant to F.M. This work was completed using resources provided by the Cambridge Tier-2 system operated by the University of Cambridge Research Computing Service (www.hpc.cam.ac.uk), funded by a EPSRC Tier-2 capital grant (EP/T022159/1). For the purpose of open access, the author has applied a Creative Commons Attribution (CC BY) license to any Author Accepted Manuscript version arising from this submission.

## SUPPLEMENTARY METHODS AND RESULTS

### 7.1 Flow Chart of Study Participant Inclusion and Exclusion

Figure S1 illustrates the flow chart detailing the inclusion and exclusion applied to study participants.

**Fig. S1.**
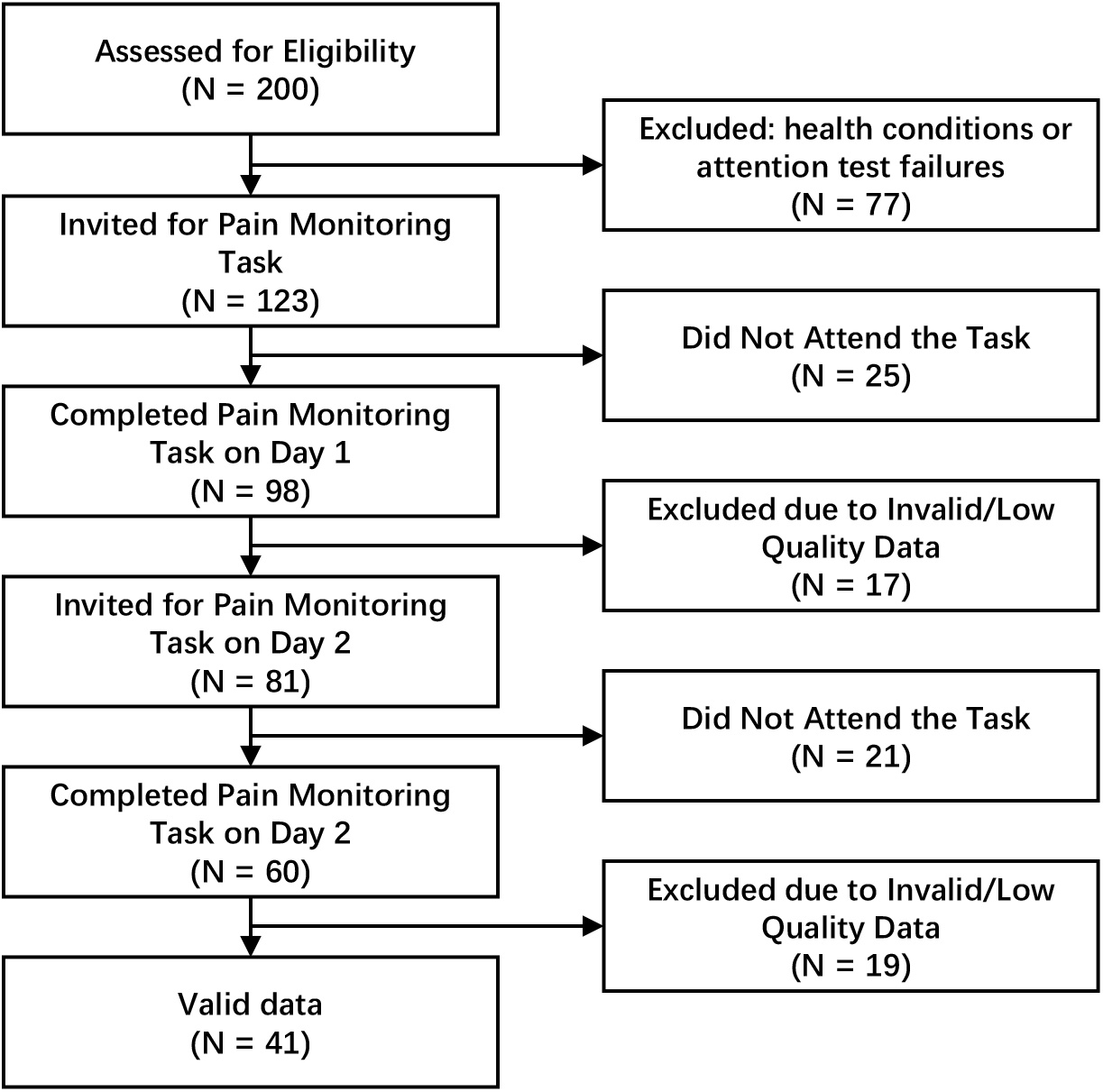
Flow Chart of Study Participant Inclusion and Exclusion. This flowchart illustrates the recruitment and screening process for the study. Of 200 participants recruited, 123 met the inclusion criteria based on health screening and attention checks. After Day 1, 81 participants completed the task, and 41 completed the two-day experiment, with exclusions due to non-attendance or invalid/low-quality data.

### 7.2 Variability analyses

Figure S2 presents the distribution of variability factors.

**Fig. S2.**
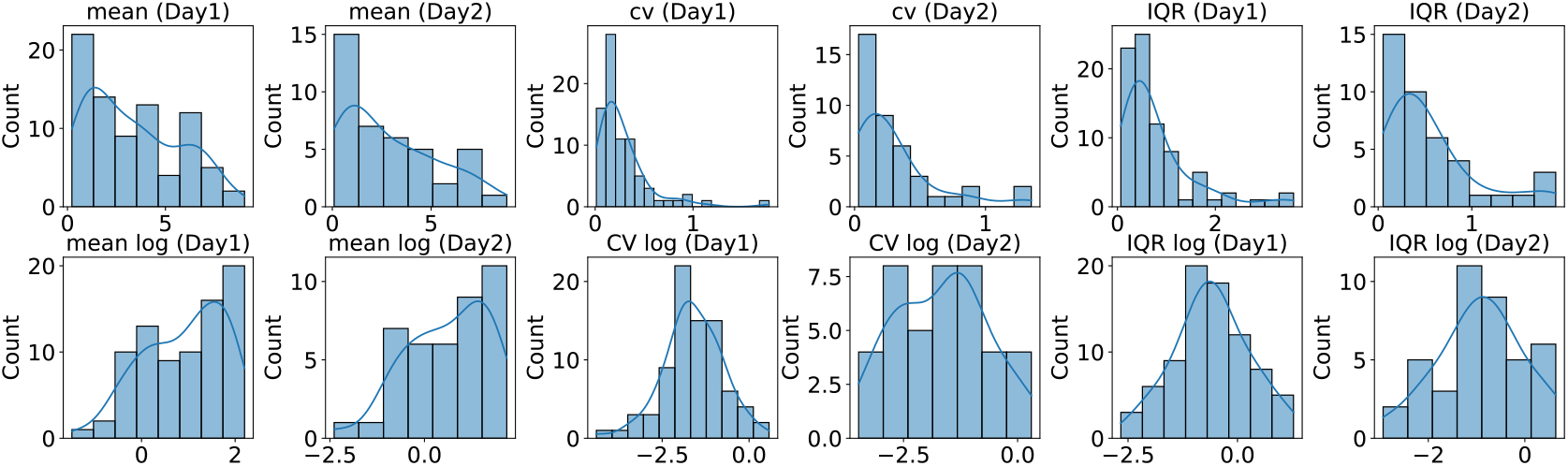
Distribution of Variability Factors Before and After Log Transformation. Distribution of variability factors (Mean, CV, IQR) for both Day 1 and Day 2 is displayed here. The first row represents the distribution based on raw data, while the second row depicts the distribution after log transformation. The distribution is less skewed after the log transformation.

Table S1 summarizes the correlations between mean pain levels, variability measures (CV, IQR), and clinical scores to explore their clinical significance.

**Table S1.** Significant Correlation between Variability Factors and Clinical Outcome (mean, CV, IQR v.s. MSK-HQ, BPI severity, PCS)

|  |  | Pearson's R | Correlation | p-value |
| --- | --- | --- | --- | --- |
| Mean (Day 1) | MSK-HQ | -0.645 | strong negative | < 0.001 |
| Mean (Day 1) | BPI Severity | 0.752 | strong positive | < 0.001 |
| Mean (Day 1) | PCS | 0.483 | moderate positive | 0.004 |
| CV (Day 1) | MSK-HQ | 0.530 | strong positive | 0.002 |
| CV (Day 1) | BPI Severity | -0.609 | strong negative | < 0.001 |
| IQR (Day 1) | BPI Severity | 0.220 |  | 0.109 |
| IQR (Day 1) | PCS | 0.299 |  | 0.061 |
| Mean (Day 2) | MSK-HQ | -0.595 | strong negative | < 0.001 |
| Mean (Day 2) | BPI Severity | 0.742 | strong positive | < 0.001 |
| Mean (Day 2) | PCS | 0.537 | strong positive | 0.001 |
| CV (Day 2) | MSK-HQ | 0.485 | moderate positive | 0.002 |
| CV (Day 2) | BPI Severity | -0.634 | strong negative | < 0.001 |
| IQR (Day 2) | BPI Severity | 0.380 | moderate positive | 0.008 |
| IQR (Day 2) | PCS | 0.445 | moderate positive | 0.003 |

Table S2 presents the correlations between variability factors and prediction accuracy.

**Table S2.**
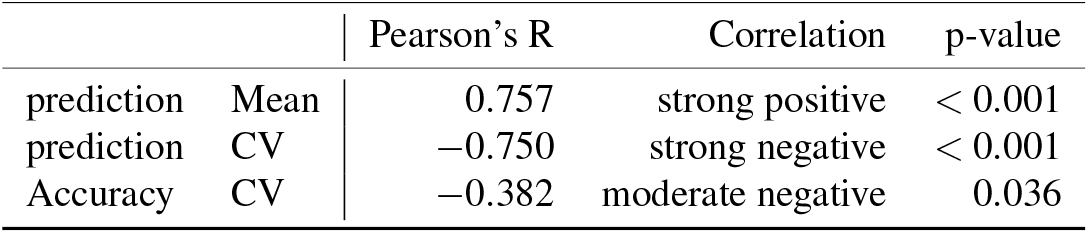
Significant Correlation between Variability Factors and Prediction Accuracy (mean, CV, IQR v.s. prediction and accuracy)

|  |  | Pearson's R | Correlation | p-value |
| --- | --- | --- | --- | --- |
| prediction | Mean | 0.757 | strong positive | < 0.001 |
| prediction | CV | -0.750 | strong negative | < 0.001 |
| Accuracy | CV | -0.382 | moderate negative | 0.036 |

## A APPENDIX

### A.1 Screening Questionnaire

1. How old are you?
2. What is your gender at birth?

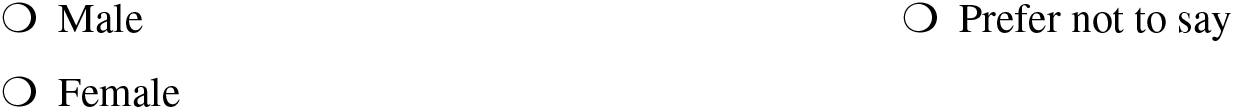
3. How long have you had pain to your **back** for?

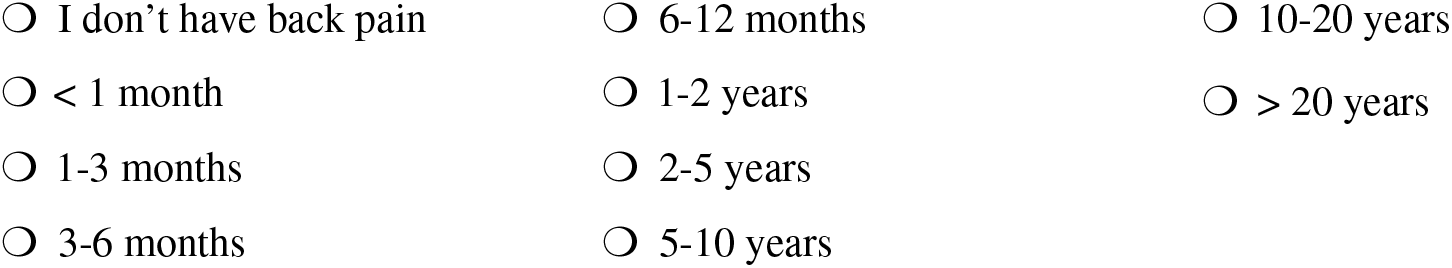
4. How long have you had pain to your **neck** for?

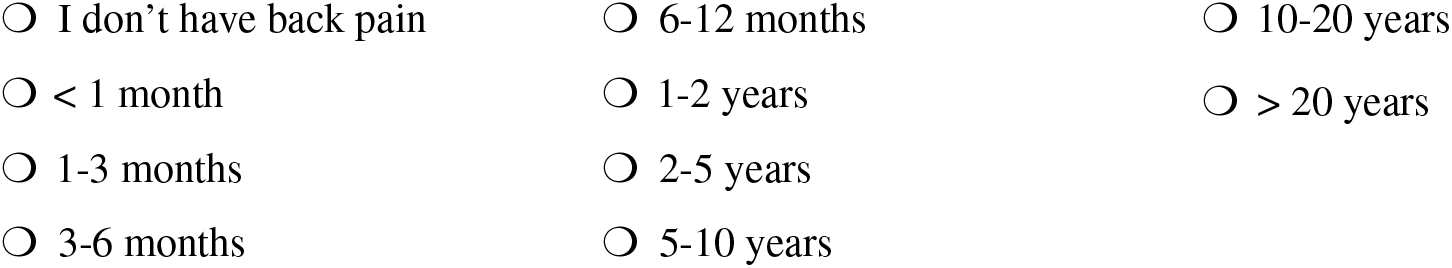
5. How long have you had pain to your **legs, knees or feet** for?

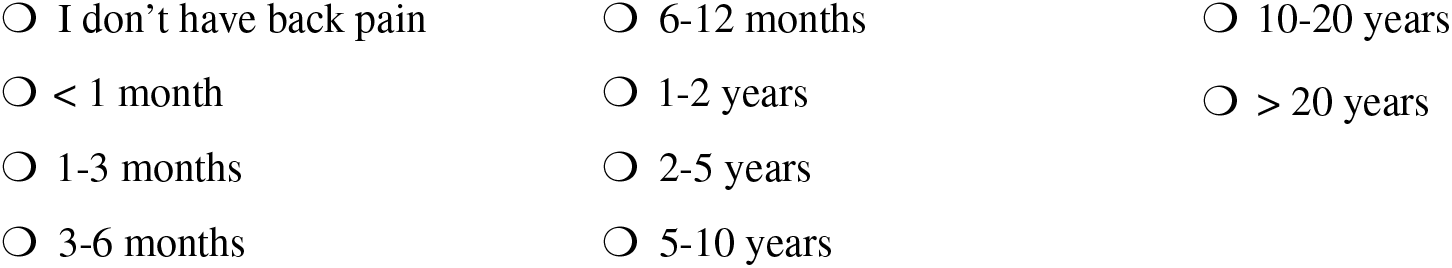
6. How long have you had pain to your **arms or hands** for?

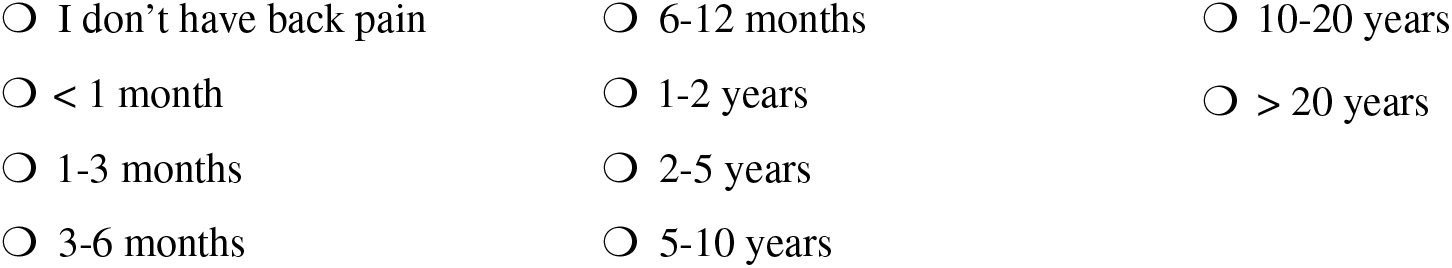
7. Do you identify as currently suffering from any of the following pain conditions? Please tick all that apply
  □ Prefer not to say
  □ Abdominal pain/IBS
  □ Arthritis (inflammatory: rheumatoid, psoriatic, etc)
  □ Arthritis (non-inflammatory, such as osteoarthritis)
  □ Complex regional pain syndrome
  □ Fibromyalgia
  □ Headache/Migraine
  □ Multiple Sclerosis
  □ Neuropathy
  □ Pelvic pain
  □ Endometriosis
  □ Trigeminal neuralgia
  □ Spinal cord injury
  □ stroke pain
  □ Other pain condition (if so, please specify below:)________
8. Have you received a diagnosis for any of these conditions? If Yes, please provide the approximate dates of diagnoses of each condition.

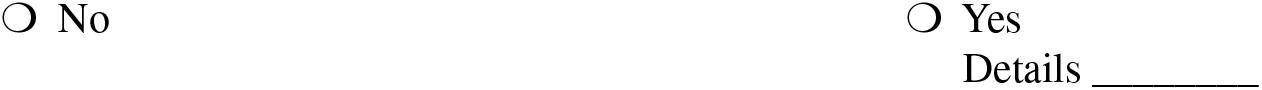
9. What treatment (pharmacological, therapy, etc), if any, are you taking for your conditions(s)? Please give detail about treatment and time periods for treatment if possible.
  - _________
10. Do you identify as living with, or have you received a diagnosis of anyother medical condition?
  ○ No
  ○ Yes If yes, please specify: Details of conditions(s) and date(s) of onset and/or diagnosis:_________
11. Do you experience any mental health difficulty? If so, how long you have experienced it, have you been diagnosed, and do you take any treatment for it? Please also provide any other details.
  - __________
12. Have you ever had or are experiencing one of the following conditions? Please tick all that apply.
  □ Amnesia (a memory difficulty)
  □ Attention deficit hyperactivity disorder (ADHD)
  □ Brain cancer
  □ Brain haemorrhage
  □ Brain injury
  □ Cognitive impairment (e.g., memory, language, attention etc)
  □ Cognitive developmental delay
  □ Dementia
  □ Dyslexia
  □ Epilepsy
  □ Hydrocephalus
  □ Learning disorder
  □ Neurodegenerative disorder
  □ Stroke
  □ Other neurological condition: Please specify:__________
  □ Other cognitive condition: Please specify:____________

